# Clinical interpretation of *KCNH2* variants using a robust PS3/BS3 functional patch clamp assay

**DOI:** 10.1101/2023.10.08.23296707

**Authors:** Kate L. Thomson, Connie Jiang, Ebony Richardson, Dominik S. Westphal, Tobias Burkard, Cordula M. Wolf, Matteo Vatta, Steven M. Harrison, Jodie Ingles, Connie R. Bezzina, Brett M. Kroncke, Jamie I. Vandenberg, Chai-Ann Ng

## Abstract

Long QT syndrome (LQTS), caused by the dysfunction of cardiac ion channels, increases the risk of sudden death in otherwise healthy young people. For many variants in LQTS genes there is insufficient evidence to make a definitive genetic diagnosis. We have established a robust functional patch clamp assay to facilitate classification of missense variants in *KCNH2*, one of the key LQTS genes. A curated set of 30 benign and 30 pathogenic missense variants were used to establish the range of normal and abnormal function. The extent to which variants reduced protein function was quantified using Z-scores; the number of standard deviations from the mean of the normalised current density of the set of benign variant controls. A Z-score of –2 defined the threshold for abnormal loss-of-function, which corresponds to 55% wild-type function. More extreme Z-scores were observed for variants with a greater loss-of-function effect. We propose that the Z-score for each variant can be used to inform the application and weighting of abnormal and normal functional evidence criteria (PS3 and BS3) within the American College of Medical Genetics and Genomics (ACMG) variant classification framework. The validity of this approach was demonstrated using a series of 18 *KCNH2* missense variants detected in a childhood onset LQTS cohort, where the level of function assessed using our assay correlated to the Schwartz score (a scoring system used to quantify the probability of a clinical diagnosis of LQTS^1^) and the length of the QTc interval.

---

Long QT syndrome (LQTS; MIM:192500) is a rare genetic disorder caused by variants in genes which encode cardiac ion channel proteins. Abnormal variants in these genes lead to cardiac ion channel dysfunction, which can result in cardiac arrhythmia and an increased risk of sudden death, often with no prior symptoms. Genetic testing is recognised as a key component of patient management and is recommended by clinical guidelines.^2^ Establishing a genetic diagnosis can inform clinical management and treatment strategies, provide greater accuracy in assessing prognosis, and enable reproductive counselling. Importantly, as not all individuals with a pathogenic variant present with clinical signs or symptoms of LQTS, where a genetic diagnosis is confirmed in an affected individual, genetic testing can be offered to family members, to identify those at risk. The perceived clinical importance of making a presymptomatic genetic diagnosis of LQTS is highlighted by the inclusion of the key causal genes in the ACMG list for reporting of secondary findings in clinical exome and genome sequencing.^3^

Current genome sequencing technologies enable more comprehensive genetic analysis to be performed cost effectively and genetic testing has become more widely adopted for LQTS. While this has led to an increase in the proportion of patients with a genetic diagnosis, the interpretation of genetic test results remains challenging. Due to lack of definitive evidence for pathogenicity, many missense variants detected in LQTS genes are classified as variants of uncertain clinical significance (VUS), which are not actionable clinically. High numbers of VUS can reduce the clinical utility of genetic testing.

In a recent study,^4^ we demonstrated the utility of a high throughput patch-clamp assay to assess the functional consequences of a set of 31 missense variants in the *KCNH2* gene (Table 1). ^4,5^ This gene encodes Kv11.1, a protein component of the voltage-gated potassium ion channel, which is responsible for the rapid delayed rectifier current (*I*_Kr_). Loss-of-function variants in this gene are one of the most common causes of LQTS.^5^ This assay, which quantifies the current density at –50mV as a surrogate for *I*_Kr_ current in heart cells, was able to clearly discriminate between missense variants with normal and abnormal loss-of-function (LoF).^4^

**Table 1:** Summary of the 60 variant controls.

| Benign/Likely Benign |  |  |
| --- | --- | --- |
| This study | 17 | Total of 30 |
| jiang *et al* 2022 | 13 |  |
| Pathogenic/Likely pathogenic |  |  |
| This study | 13 | Total of 30 |
| jiang *et al* 2022 | 17 |  |

The details for each of the variant controls are available in Table S1.

We previously illustrated how data from this assay could be used to inform clinical variant classification using the American College of Medical Genetics and the Association for Molecular Pathology (ACMG/AMP) framework, providing statistical support for application of the functional evidence (PS3 and BS3 criteria) with ‘supporting’ or ‘moderate’ weighting.^4^ In this study, we expand on our previous work,^4^ undertaking functional analysis of additional variants to give a total of 30 pathogenic and 30 benign variant controls (Table 1). The 30 benign / likely benign variant controls were selected on the basis of their minor allele frequency (MAF) in the Genome Aggregation database (gnomAD;maximum population filtering allele frequency^6^ [PopMax FAF] greater than 5x10^−5^, see supplementary materials). The advantage of using a larger number of benign variant controls, selected based on the MAF, is that we were able to sample a broader range of biological and experimental variation, and more clearly define a range for normal function measured by patch clamp electrophysiology (Figure 1 and Figure S2). Using our assay, the functionally normal range was defined by a Z-score within ±2 of the mean normalised current density determined from 30 benign control variants. The functionally abnormal LoF range was defined by a Z-score of –2 or below, corresponding to a variant having less than 55% of normal function, a clinically relevant threshold consistent with protein haploinsufficiency.^7^

**Figure 1:**
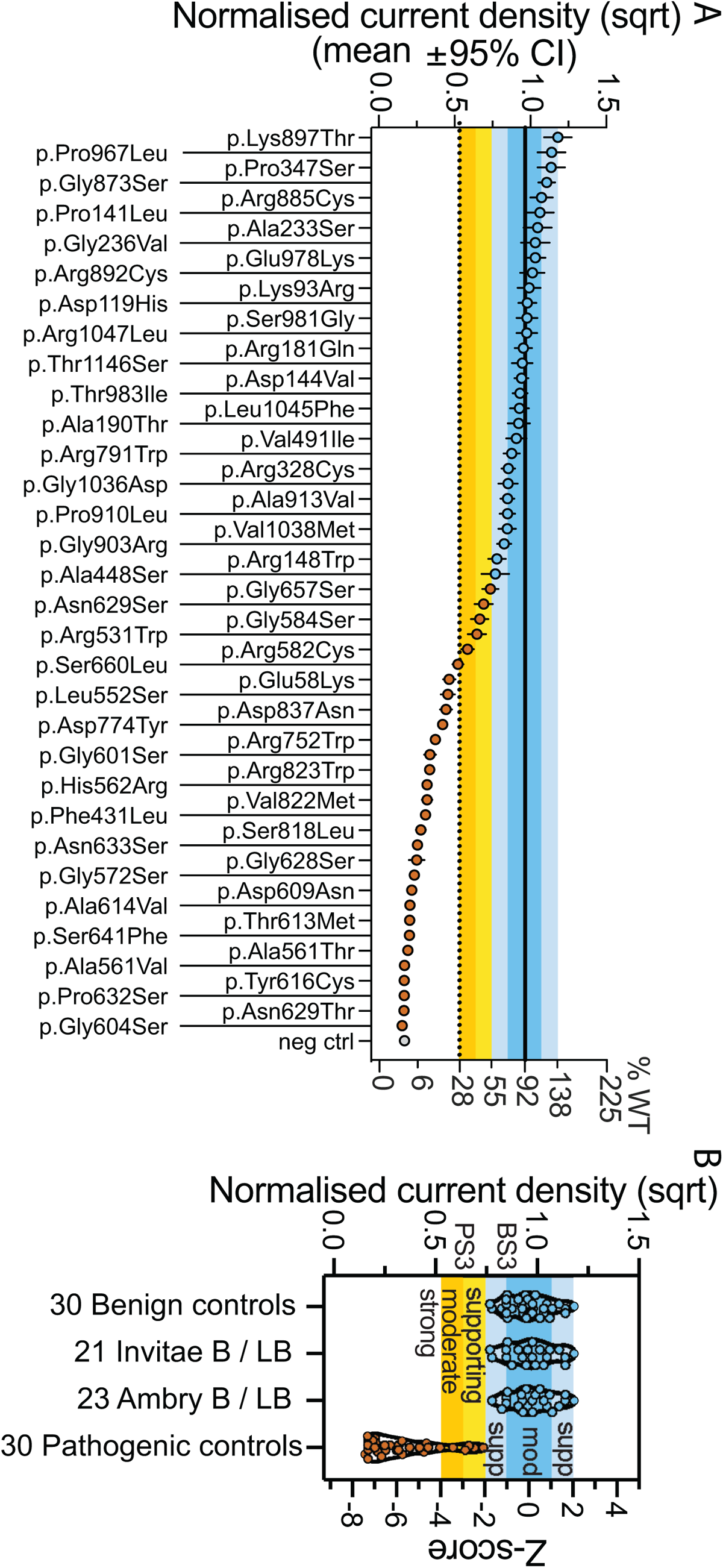
Establishment of graded functional evidence strength based on the Z-score determined from benign and pathogenic variant controls. (A) The cut-off for functionally normal (blue circles) and abnormal (brown circles) was determined to be 55% of WT using 2 standard deviations below the mean of all benign controls. (B) The establishment of evidence strength for functional data based on the Z-score of –2 by using 30 benign variant controls. The level of evidence strength was graded based on the Z-score where normal function for BS3 at moderate (between 0 and ± 1) or supporting (between 1 and 2 or –1 and –2). The abnormal function for PS3 at supporting (between –2 to –3), moderate (between –3 to –4) and strong (less than –4). The comparison of benign variant controls was made against known benign / likely benign variants classified by two other diagnostic laboratories. All the 30 pathogenic variant controls used were found to have function less than the benign variant controls (see Table S3).

In our previous study, the 17 pathogenic variant controls had various levels of reduced function. In this study, we included an additional 13 pathogenic variant controls (Table 1 and Table S1) to increase the statistical power of the assay (Table 2). The experimental procedure is described in the supplementary materials. Four out of the 30 pathogenic variant controls were observed in gnomAD, however, with the exception of the Finnish founder variant, p.(Leu552Ser),^7,8^ all had a minor filtering allele frequency lower than the threshold selected for benign control variants (Table S1). All 30 pathogenic variant controls had Z-scores of –2 or less (Figure 1 and Table S3). Assay performance was evaluated using the approach recommended by the Clinical Genome Resource (ClinGen) Sequence Variant Interpretation (SVI) Working Group^9^; the odds of pathogenicity (OddsPath) were determined to be 30 for abnormal function (PS3) and 0.034 for normal BS3 function (Table 2).

**Table 2:**
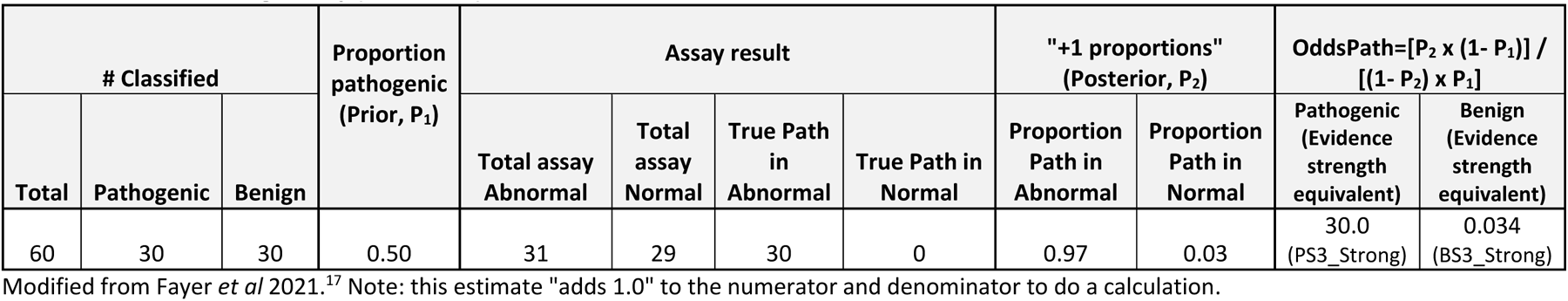
Odds of Pathogenicity (OddsPath).

To integrate this functional data into variant classification, and to take into consideration the various levels of reduced function revealed by this assay, we considered a graded evidence scale weighted according to the severity of the LoF determined by the Z-score. Using this scale, variants with Z-score between –2 and –3 (mild LoF), –3 and –4 (moderate LoF) and less than –4 (severe LoF) could be categorised as PS3_supporting, PS3_moderate or PS3_strong, respectively. It is important to note that this *in vitro* cell model system may not fully reflect protein function *in vivo* and might not capture all abnormal LoF. Taking into consideration these limitations, we proposed that normal function variants with a Z-score between 0 and 1 or –1 could be classified as BS3_moderate and Z-score between 1 and 2 or –1 and –2 as BS3_supporting (Figure 1).

To investigate the utility of our approach, the functional evidence level determined from this assay was applied to enhance the classification of 18 *KCNH2* missense variants detected by Westphal *et al* in children with suspected LQTS.^10^ Evidence of abnormal LoF was found for 14 missense variants (Figure 2A, yellow and brown circles and Table S3). Four variants were in the range of normal function (Table S3). Two of these variants (p.Asp259Asn and p.Thr983Ile), were detected in children who had been included in the cohort due to a family history sudden cardiac death, but had normal QTc and a Schwartz score of less than 1 (low probability of LQTS^1^) (Figure 2C). The p.Val409Ile variant was found in a child with a Schwartz score of 2 (intermediate probability of LQTS^1^). The p.Pro393Leu variant was found in a child who was also heterozygous for the p.Arg176Trp variant, a possible risk allele,^11^ which has been described as a founder variant in the Finnish population.^12^

**Figure 2:**
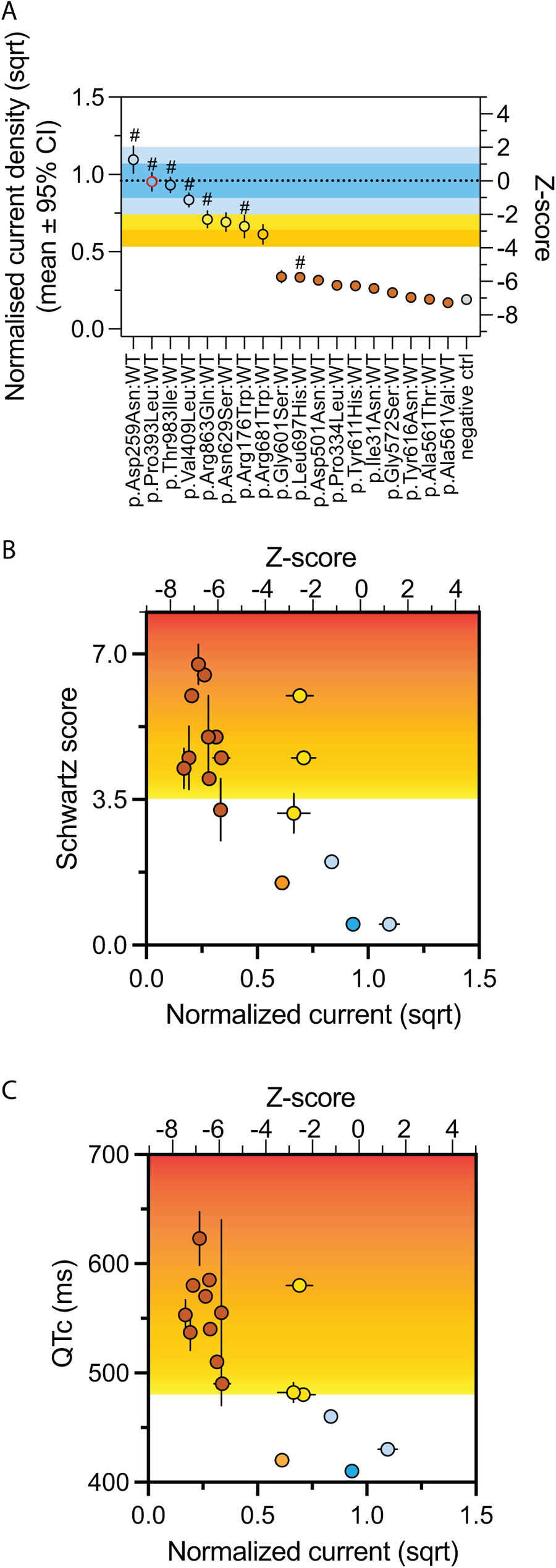
Missense *KCNH2* variants from children with long QT syndrome. (A) The functional assessment of these variants using patch-clamp assay. Blue region indicates the functionally normal range determined from a set of 30 benign variant controls. Variants are colour-coded to indicate the Z-score: blue for Z-score between 0 and ±1, light blue for Z-score between 1 and 2 or –1 and –2, yellow for Z-score between –2 to –3, orange for Z-score between –3 to –4 and brown for Z-score less than – 4. Variants that were downgraded to VUS in Westphal *et al*^10^ are indicated by #. The heterozygous variant p.(Pro393Leu) with a risk allele p.(Arg176Trp) is indicated by red outline. (B) The correlation of Schwartz score from children that have these variants and the corresponding function of the variant assessed by patch clamp assay. Region of Schwartz score above 3.5 is highlighted to indicate the severity of the LQT clinical phenotype. The Pearson r for this correlation is –0.74 and the R^2^ of linear regression is 0.54 with p=0.0007. The Schwartz score for the compound heterozygous p.(Pro393Leu) with a risk allele p.(Arg176Trp) was not included. Data is presented as mean +/- s.e.m. (C) The correlation of QTc and the corresponding function of the variant assessed by patch clamp assay. Region of QTc above 480ms is highlighted. The Pearson r for this correlation is –0.76 and the R^2^ of linear regression is 0.58 with p=0.0004. The QTc for the compound heterozygous p.(Pro393Leu) with a risk allele p.(Arg176Trp) was not included. Data is presented as mean +/- s.e.m.

Considering data from 32 paediatric LQTS cases in this cohort who were heterozygous for one of these *KCNH2* missense variants, there was a negative correlation between the Z-score determined by this assay and Schwartz score (r = –0.74 (Figure 2B) with a R2 of 0.54 from linear regression). Overall, the variants that were determined by our assay to have severe LoF were detected in individuals with a more severe clinical phenotype (measured by Schwartz score) and a more prolonged QTc, this is consistent with previous observations.^13,14^ The variant classifications in Westphal *et al*^10^ were revised by applying the level of functional evidence at the strength determined by the Z-scores (Table S3). Considering this additional evidence, the classification of one variant (p.Leu697His) was upgraded from VUS to likely pathogenic after PS3 was applied at strong level (Table S2).

Overall, the findings of our study indicate that the functional evidence produced by this validated assay is extremely valuable for the interpretation of *KCNH2* missense variants in the context of LQTS. However, functional data from this assay should not be used as sole evidence to classify a variant as pathogenic or benign, but should be considered in the context of other evidence, as outlined in the ACMG/AMP framework.^15^

At present, we have not extensively examined the functional range of so-called risk alleles, which display incomplete penetrance and may only have pathogenic potential in the presence of other environmental or genetic risk factors. For example, the p.(Arg176Trp) variant, which was detected in the Westphal *et al* childhood onset LQTS cohort, has been reported as founder variant in the Finnish population^8^ and reported to be enriched in individuals referred for LQTS genetic testing but lacking a molecular diagnosis.^16^ The frequency of this variant in the gnomAD European (non-Finnish) population was higher than the threshold used to select benign variant controls for this study, however, it was determined by the assay to have a mild LoF effect (Z-score –2.75); these findings are consistent with this being a variant with low or incomplete penetrance. Further studies may provide additional functional evidence which might help clarify the pathogenic potential of this class of variant.

In conclusion, this patch clamp assay, which used extensively characterised variant controls to calibrate thresholds for normal and abnormal loss-of-function missense variants in the *KCNH2* gene, provides reliable functional evidence that can be incorporated into clinical variant classification. This has potential to reduce uncertainty in variant interpretation in this clinically important gene.

## Data and code availability

The current traces from the patch clamp electrophysiology that support the findings of this study are available as csv files upon request. The corresponding analysis scripts for patch-clamp experiments are available at https://git.victorchang.edu.au/projects/SADA/repos/syncropatch_automated_analysis/browse.

## Acknowledgements

This work was funded by an NSW Cardiovascular Disease Senior Scientist grant (J.I.V.), an National Health and Medical Research Council Principal Research Fellowship (J.I.V.), MRFF Genomics Health Futures Mission grant (J.I.V. and C.A.N). We also acknowledge support from the Victor Chang Cardiac Research Institute Innovation Centre, funded by the NSW Government.

## Author contributions

Conceptualization: K.L.T., J.I.V., C.A.N.; Data curation: K.L.T., E.R., C.A.N.; Formal Analysis: K.L.T., C.J., E.R., D.S.W., T.B., C.M.W., M.V., S.M.H., J.I.V., C.A.N.; Funding acquisition: J.I.V.; Methodology: K.L.T., C.J., E.R., J.I., J.I.V., C.A.N.; Writing – original draft: K.L.T., J.I.V., C.A.N.; Writing – review & editing: K.L.T., E.R., D.S.W., T.B., C.M.W., M.V., S.M.H., J.I., C.R.B., B.M.K., J.I.V., C.A.N.

