## supplemental materials for "Clinical interpretation of *KCNH2* variants using a robust PS3/BS3 functional patch clamp assay"

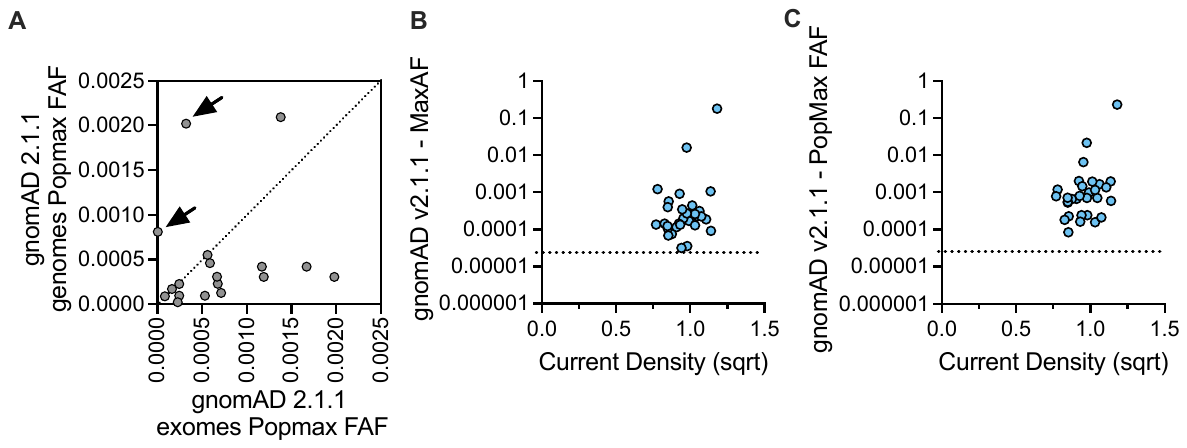

**Figure S1: The MaxAF and PopMax FAF of benign variant controls.** (A) Comparison between the PopMax FAF from the exomes and genomes of gnomAD v2.1.1. In general, the PopMax FAF from the exomes is expected to be more reliable than the genomes. Arrows indicate the two outliers (p.Ala190Thr and p.Leu1045Phe). (B) The plot of MaxAF from gnomAD v2.1.1 and corresponding current density for benign variant controls. (C) The plot of PopMax FAF from gnomAD v2.1.1 and corresponding current density for benign variant controls. See Table S1 for more details.

**Supplemental Methods**

**Selection of benign variants control**

Benign and likely benign *KCNH2* missense variants were curated using data from the gnomAD database (Table S1) by identifying *KCNH2* variants which would be considered too common in gnomAD to be causal of monogenic LQTS, a maximum credible population allele frequency (MaxAF) was calculated using the approach described by Whiffin *et al*.^1^ This MaxAF was calculated using the allele frequency app available at <https://www.cardiodb.org/allelefrequencyapp/>. This calculation considered a disease prevalence estimate of 1 in 2,000;^2^ a maximum allelic contribution of 2% (taking into consideration pathogenic *KCNH2* variant frequency from two large LQTS patient cohorts^3^ and OMGL); and a conservative estimate of variant penetrance of 10%. See Table S4 for further details. *KCNH2* missense variants with gnomAD PopMax filtering allele frequency (FAF) greater than our calculated MaxAF (5x 10^-5^) was considered sufficient evidence to allow application of the benign strong ACMG criteria, BS1, and in the absence of any conflicting data, to support a ‘Likely’ Benign classification. Typically, the FAF from the exomes in v2.1.1 is more reliable than the genomes due to greater allele count but in the event where PopMax FAF is not available (p.Gly236Val) or not reliable from the exomes in v2.1.1 (p.Ala190Thr and p.Leu1045Phe) (Figure S1A), the PopMax FAF from the genomes in v2.1.1 was used instead (Table S1). A total of 30 *KCNH2* missense variants in gnomAD that met this criterion was selected as benign variant controls (Figure S1B and C).

**Selection of pathogenic variants control**

A total of 30 *KCNH2* missense pathogenic variant controls were available in Table S1. The set of 17 *KCNH2* missense pathogenic variant controls used in the Jiang *et al*^4^ was supplemented with an additional 13 missense variants in this study, which were selected based on multiple submitters in ClinVar as likely pathogenic / pathogenic, as well as by the Oxford Genetics Laboratories, to ensure all the abnormal range have been properly sampled.

**The automated patch clamp assay**

A method protocol describing the DNA plasmids, generation of heterozygous *KCNH2* Flp-In HEK293 cell lines, cell lines maintenance and cell harvesting for automated patch clamp assay was published previously.^5^ Briefly, DNA plasmids were generated by GenScript Inc. (Pistcataway, NJ, USA) to generate Flp-In HEK293 (Thermo Fisher, cat. #R78007) stable cell lines, which were assayed using an APC electrophysiology platform (SyncroPatch 384PE, Nanion Technologies, Munich, Germany). The functionally normal and abnormal range was defined using the cut-off of Z-scores at ±2. The ‘Odds of Pathogenicity’ were calculated based on the recommendation by the ClinGen SVI Working Group.^6^
